# Exploring the extent of uncatalogued genetic variation in antimicrobial resistance gene families in *Escherichia coli*

**DOI:** 10.1101/2023.03.14.23287259

**Authors:** Samuel Lipworth, Derrick Crook, A. Sarah Walker, Tim Peto, Nicole Stoesser

## Abstract

**Background:** Antimicrobial resistance (AMR) in *E. coli* is a global problem associated with substantial morbidity and mortality. AMR-associated genes are typically annotated based on similarity to a variants in a curated reference database with an implicit assumption that uncatalogued genetic variation within these is phenotypically unimportant. In this study we evaluated the potential for discovering new AMR-associated gene families and characterising variation within existing ones to improve genotype-to-susceptibility-phenotype prediction in *E. coli*.

**Methods:** We assembled a global dataset of 9001 *E. coli* sequences of which 8586 had linked antibiotic susceptibility data. Raw reads were assembled using Shovill and AMR genes extracted using the NCBI AMRFinder tool. Mash was used to calculate the similarity between extracted genes using Jaccard distances. We empirically reclustered extracted gene sequences into AMR-associated gene families (70% match) and alleles (ARGs, 100% match).

**Results:** The performance of the AMRFinder database for genotype-to-phenotype predictions using strict 100% identity and coverage thresholds did not meet FDA thresholds for any of the eight antibiotics evaluated. Relaxing filters to default settings improved sensitivity with a specificity cost. For all antibiotics, a small number of genes explained most resistance although a proportion could not be explained by known ARGs; this ranged from 75.1% for co-amoxiclav to 3.4% for ciprofloxacin. Only 17,177/36,637 (47%) of ARGs detected had a 100% identity and coverage match in the AMRFinder database. After empirically reclassifying genes at 100% nucleotide sequence identity, we identified 1292 unique ARGs of which 158 (12%) were present ≥10 times, 374 (29%) were present 2-9 times and 760 (59%) only once. Simulated accumulation curves revealed that discovery of new (100%-match) ARGs present more than once in the dataset plateaued relatively quickly whereas new singleton ARGs were discovered even after many thousands of isolates had been included. We identified a strong correlation (Spearman coefficient 0.76 (95% CI 0.72-0.79, p<0.001)) between the number of times an ARG was observed in Oxfordshire and the number of times it was seen internationally, with ARGs that were observed 7 times in Oxfordshire always being found elsewhere. Finally, using the example of *bla*_*TEM-1*_, we demonstrated that uncatalogued variation, including synonymous variation, is associated with potentially important phenotypic differences (e.g. two common, uncatalogued *bla*_*TEM-1*_ alleles with only synonymous mutations compared to the known reference were associated with reduced resistance to co-amoxiclav [aOR 0.57, 95%CI 0.34-0.93, p=0.03] and piperacillin-tazobactam [aOR 0.54, 95%CI 0.32-0.87, p=0.01]).

**Conclusions:** Overall we highlight substantial uncatalogued genetic variation with respect to known ARGs, although a relatively small proportion of these alleles are repeatedly observed in a large international dataset suggesting strong selection pressures. The current approach of using fuzzy matching for ARG detection, ignoring the unknown effects of uncatalogued variation, is unlikely to be acceptable for future clinical deployment. The association of synonymous mutations with potentially important phenotypic differences suggests that relying solely on amino acid-based gene detection to predict resistance is unlikely to be sufficient. Finally, the inability to explain all resistance using existing knowledge highlights the importance of new target gene discovery.

## INTRODUCTION

Antimicrobial resistance (AMR) is a major global challenge with substantial associated morbidity and mortality[1]. In E. coli, AMR is mostly conferred by acquisition of genes which can be integrated into the chromosome or carried on plasmids[2–4]. It can also occur via point mutations in both core and accessory genes. Extensive efforts to catalogue and characterise these mechanisms have been undertaken, resulting in several highly curated databases which are widely used for genomic epidemiology[5–7].

Several studies have investigated the performance of such databases to predict phenotype from genotype for *E. coli*[8, 9], demonstrating the associated challenges. Verschuuren et al. recently demonstrated the inability of the Resfinder tool to meet FDA specifications for very major/major error rates for most antibiotics (n=234 isolates, selected for resistance to third-generation cephalosporins[10]). This work highlighted particularly poor performance predicting AMR phenotype for beta lactam beta-lactamase inhibitor (BL-BLI) combination drugs, replicating a finding from earlier studies[11, 12]. Feldgarden et al claimed much better performance (99.7% overall concordance when pooling across antibiotic classes) albeit with a small (n=47 isolates) and predominantly susceptible dataset[6]. These and other studies highlight the need for further development and expansion of these databases if they are to become clinically useful.

There are two hierarchical levels of annotation within existing AMR gene catalogues: AMR-associated gene families (e.g. *bla*_*CTX-M*_, *gyrA*) and alleles of gene families (e.g. *gyrA S*83*L, bla*_*CTM-M-15*_, *bla*_*CTX-M-27*_) which we hereby refer to as antibiotic resistance genes (ARGs). It is currently standard practice to characterise the presence or absence of ARGs based on percentage identity and coverage (e.g. commonly used thresholds for the former are 80% [the ABRicate default[13]] and 90% [the AMRFinder default[13]]). Where there is no perfect match, the presence of the closest characterised allele in the same gene family is reported and hence most molecular epidemiology and resistance prediction studies currently ignore any non-catalogued variation. To our knowledge the prevalence, diversity and impact of these imperfectly matching genes has not been systematically evaluated. Improvements to existing catalogues may therefore come either from discovery of novel AMR-associated gene families and/or improved annotation of variation within existing ones.

In this work we seek to estimate the potential for further exploration of these two domains of genomic variation to improve existing ARG reference databases. We firstly quantify how much resistance is explained by presence or absence of known ARGs/variants and therefore estimate how much might be gained by searching for novel AMR-associated gene families. Secondly we explore variation within known AMR-associated gene families at 100% match (acknowledging that this may be caused by sequencing/assembly error as well as true biological variation) which is currently un-catalogued and investigate whether including this in future versions of databases is likely to be useful.

## METHODS

### Isolates and Assembly

Five large E. coli sequencing projects (predominantly from human bloodstream infections: PRJEB11403[14], PRJEB2329412, PRJEB32059[15], PRJEB4681[16] and PRJNA604975[17]) were selected for inclusion because they had linked whole genome sequencing and antimicrobial susceptibility phenotype data available. These data represented distinct geographic regions, including Thailand, Norway, Sweden, Australia and the UK. Raw reads from N=9001 isolates in these BioProjects were downloaded from the ENA and subsequently assembled using Shovill (v1.0.4[18]) with default settings. Of these, N=8,586 (95.4%) isolates had associated antimicrobial susceptibility data (all binary and measured using EUCAST breakpoints) for at least one antibiotic. Details of all isolates included and assembly statistics are available in supplementary file S1.

### Gene sequence extraction

We chose to focus on ARGs encoding resistance to eight drugs in six clinically relevant antibiotic classes (as defined by AMRFinder) for the treatment of E. coli infection in humans, namely: aminoglycosides (gentamicin), beta-lactams (ampicillin and the beta-lactamase inhibitor combinations co-amoxiclav and piperacillin-tazobactam), cephalosporins (ceftriaxone), fosfomycin, quinolones (ciprofloxacin) and trimethoprim. Other ARGs were excluded from this analysis. We ran the AMRFinder software (v3.10.23[19], database version 2022-12-19.1) using the -O Escherichia --nucleotide output flags (using the default curated or 90% identity threshold and default 50% minimum coverage threshold). We then extracted all sequences for each antibiotic class into a single multi-fasta file (e.g. aminoglycoside.fasta, beta lactam.fasta). We then sketched these sequences (Mash sketch -s 100,000 -i) and created an all vs all distance matrix from which we calculated distance as the number of shared hashes divided by the total number of hashes.

Given that AMR gene nomenclature sometimes assigns similar gene names to ARGs that are genetically diverse, and different gene names to ARGs that are genetically similar, we empirically redefined AMR-associated gene families and ARGs (Figure 1). AMR associated gene families were defined by filtering mash distance matrices for any given antibiotic class at a ≥0.7 similarity threshold (e.g. 70% kmer of all possible kmers match exactly, approximately similar to the threshold used by e.g. Panaroo to define gene families). No such reclassification was performed for genes belonging to the “point” AMRFinder element subtype (e.g. *gyrA, parC*) as these are core genes that are not difficult to accurately identify. These were then converted into graphs from which communities (AMR-associated gene families) were detected using complete linkage (R package igraph[20]). AMR-associated gene families were named according to the most common label assigned to their members by AMRFinder. We hereby refer to each unique version (including the reference sequence/wild-type) of any AMR-associated gene family as an ARG, regardless of whether it contains one or more SNPs/indels compared to the reference sequence. To define these ARGs we repeated the process above with a 1.0 similarity threshold (i.e. 100% sequence identity and coverage) and assigned sequential numeric labels to define unique ARGs within a given AMR-associated gene family (e.g. *bla*_*TEM-1 1*_, *bla*_*TEM-1 2*_, etc).

**Figure 1.**
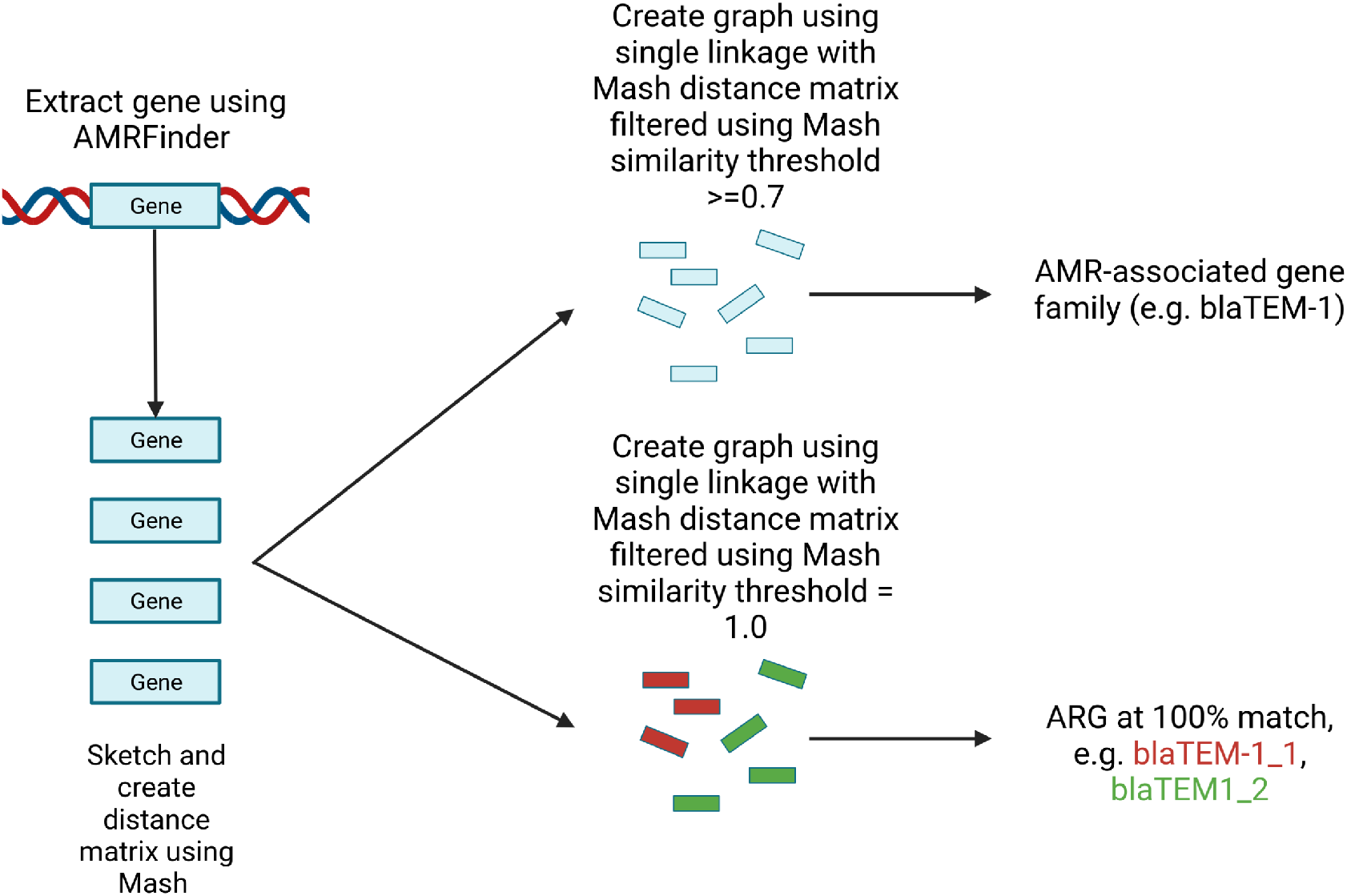
Method used for empirical definition of AMR-associated gene families (clusters of genes identified at 70% similarity threshold) and ARGs (clusters of genes identified at 100% similarity threshold).

### Statistical analysis

Isolates were predicted as being resistant to an antibiotic if their genotype contained any allele associated with resistance to the drug in the AMRFinder database (this analysis was first performed with strict 100% coverage and identity filters and subsequently using the AMRFinder default settings [i.e. 90% or curated identity threshold, 50% coverage]). Because AMRFinder does not provide any phenotypic sub-classifications of beta-lactam-resistance encoding ARGs, we used the lookup table provided by ResFinder[5] to predict phenotype for the beta-lactam/beta-lactamase inhibitor drugs. Sensitivity, specificity, negative predictive value (NPV), positive predictive value (PPV) were calculated in the standard way, making comparisons against the laboratory-derived antibiotic susceptibility phenotypes that were available for the sequences analysed as the reference standard.

We also calculated the frequency of “major discrepancy” (“maj”; i.e. erroneous genotypic prediction of susceptible isolates as resistant when compared with the reference phenotype) and very major discrepancy (“vmj”; i.e. erroneous genotypic prediction of resistant isolates as susceptible when compared with the reference phenotype) in line with the US FDA’s guidance on acceptable performance standards for antimicrobial susceptibility test systems, where major discrepancies need to be <3% and are defined as: (Number maj discrepancies / Total number of susceptible organisms by reference method) X 100 and very major discrepancies are defined as: (Number vmj discrepancies / Total number of resistant organisms by reference method) X 100). This is dependent on the number of resistant organisms tested, with an acceptable upper 95% confidence limit for the true vmj rate set at <7.5%[21].

We estimated exact binomial 95% confidence intervals using the R package Stats. Rarefaction curves were created after randomisation of the isolate order using the “rarefaction” function of R package Micropan[22] (n.perm=100). Firth regression (R package logistf[23]) was used to investigate whether different 100% match variants of *bla*_*TEM-1*_ were associated with a higher probability of resistance to co-amoxiclav/piperacillin-tazobactam.

### Checks for possible sequencing/bioinformatic error

To investigate the extent to which sequencing/bioinformatic error might inflate the true number of ARGs that occurred only once (“singletons”) we firstly evaluated the median read depth in singleton vs non-singleton ARGs. For each isolate, we used Snippy (v4.6.0[24]) to map raw reads back to the FASTA file created by the AMRFinder --nucleotide-output option. We then used samtools depth -a to evaluate the depth at every position in all ARGs and then summarised this output to give a median depth for each ARG. A Wilcoxon Rank Sum test was used to compare the distributions of read depths in singleton vs non-singleton ARGs. The assumption was that variants generated through sequencing error may occur due to lower read depth. We also selected 1000 random isolates (sort -R all isolates | tail -1000) with at least 1 singleton ARG and reassembled these using SKESA[25] (v.2.4.0) to evaluate the proportion of singleton ARGs that were identically assembled using two independent assembly methods (i.e. Shovill and SKESA).

### Data availability

All assemblies are available at 10.6084/m9.figshare.22220212 and associated metadata can be found in supplementary dataset 1. All code used for the analysis can be found at https://github.com/samlipworth/resistome variation where there is also a binder environment in which the key aspects of the analysis can be replicated.

## RESULTS

### Not all phenotypic antimicrobial resistance can be explained using the genotypic catalogue represented in the current AMRFinder database

We assembled a collection of 8,586 E. coli isolates with linked whole genome sequencing data and binary phenotypic classifications available for at least one antibiotic of interest (details in Table S1). The sensitivity of the AMRFinder database (i.e. percentage of phenotypically resistant isolates with a relevant ARG as determined by AMRFinder) using 100% identity/coverage filters was notably poorer for beta-lactam/beta-lactamase inhibitor (BL-BLI) combinations (co-amoxiclav 24.9% (95% CI 22.7-26.9), piperacillin-tazobactam 40.7% (35.3-45.9) and fosfomycin (6.7% (0.3-44.0)) than other antibiotics considered (Table 1). The sensitivity for both co-amoxiclav and piperacillin-tazobactam could be improved by predicting isolates carrying blaTEM-1 as resistant but this reduced specificity (increase in sensitivity: 61.5%/47.1% but reduced specificity: 29.5%/36.1% for co-amoxiclav/piperacillin-tazobactam respectively). Using 100% identity/coverage filters no drugs met the FDA specified acceptable thresholds for very major discrepancy rates but seven (gentamicin, ceftriaxone, ciprofloxacin, co-amoxiclav, ampicillin, fosfomycin and trimethoprim) met the acceptable thresholds for major discrepancy rates (Table 1). When identity/coverage settings were relaxed to default settings, there was a increase in sensitivity but drop in specificity for six drugs (gentamicin, ampicillin, ciprofloxacin, ceftriaxone, fosfomycin and trimethoprim). This increased the major discrepancy rates for ciprofloxacin, ceftriaxone and fosfomycin to above the FDA specified acceptable threshold (Table 1).

**Table 1.**
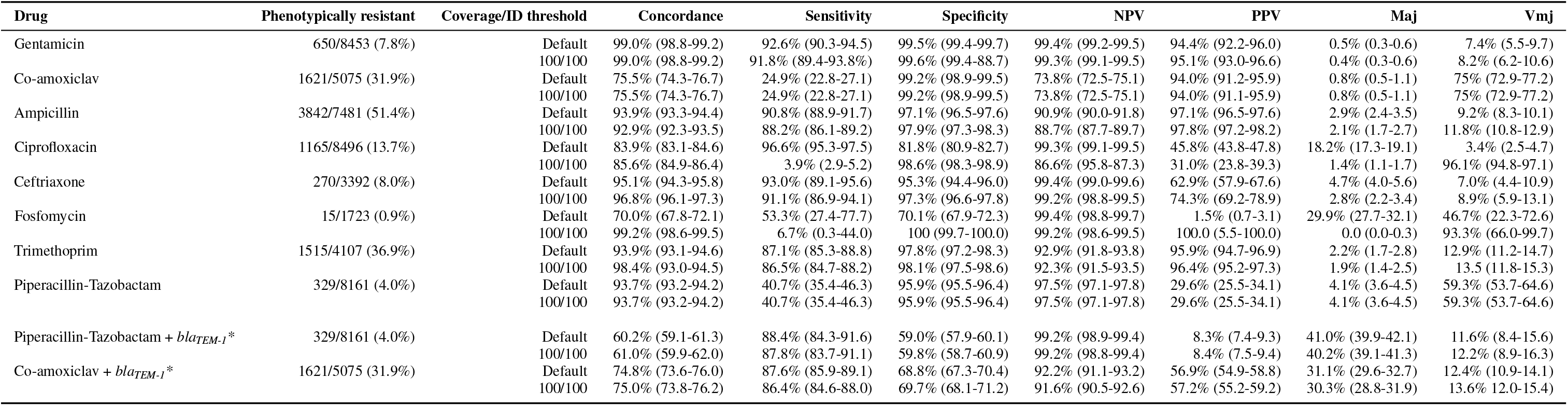
Performance metrics for the ability of the AMRFinder database to predict phenotype from genotype. Performance metrics that did not meet the FDA thresholds for major discrepancies (Maj - <3%) or very major discrepancies (Vmj upper bound of 95% confidence interval <7.5%) are highlighted in blue. NPV - negative predictive value, PPV - positive predictive value. Values in parenthesis represent 95% confidence intervals. Coverage/ID threshold refers to the % amino acid coverage/identity of the gene identified by AMRFinder compared to the reference. *in these rows blaTEM is classified as conferring resistance against co-amoxiclav/piperacillin-tazobactam respectively.

For all antibiotic classes, the majority of explainable resistance was conferred by a small number of ARGs/mutations, with a large number of rarer alleles contributing relatively little (Figure 2). The proportion of resistant isolates was particularly variable for some alleles/ARGs classified by AMRFinder as conferring resistance to ciprofloxacin, fosfomycin, piperacillin-tazobactam and trimethoprim. The greatest phenotypic heterogeneity occurred with carriage of blaTEM-1 for co-amoxiclav (1107/2130 52.0% isolates resistant) and piperacillin-tazobactam (197/3099 6.4% isolates resistant). All antibiotics had a sensitivity “gap”, i.e. a proportion of resistance which could not be explained using all ARGs/mutations included in the current AMRFinder catalogue (at the default identity thresholds), but this varied by drug, from 75.1% for co-amoxiclav to 3.4% for ciprofloxacin.

**Figure 2.**
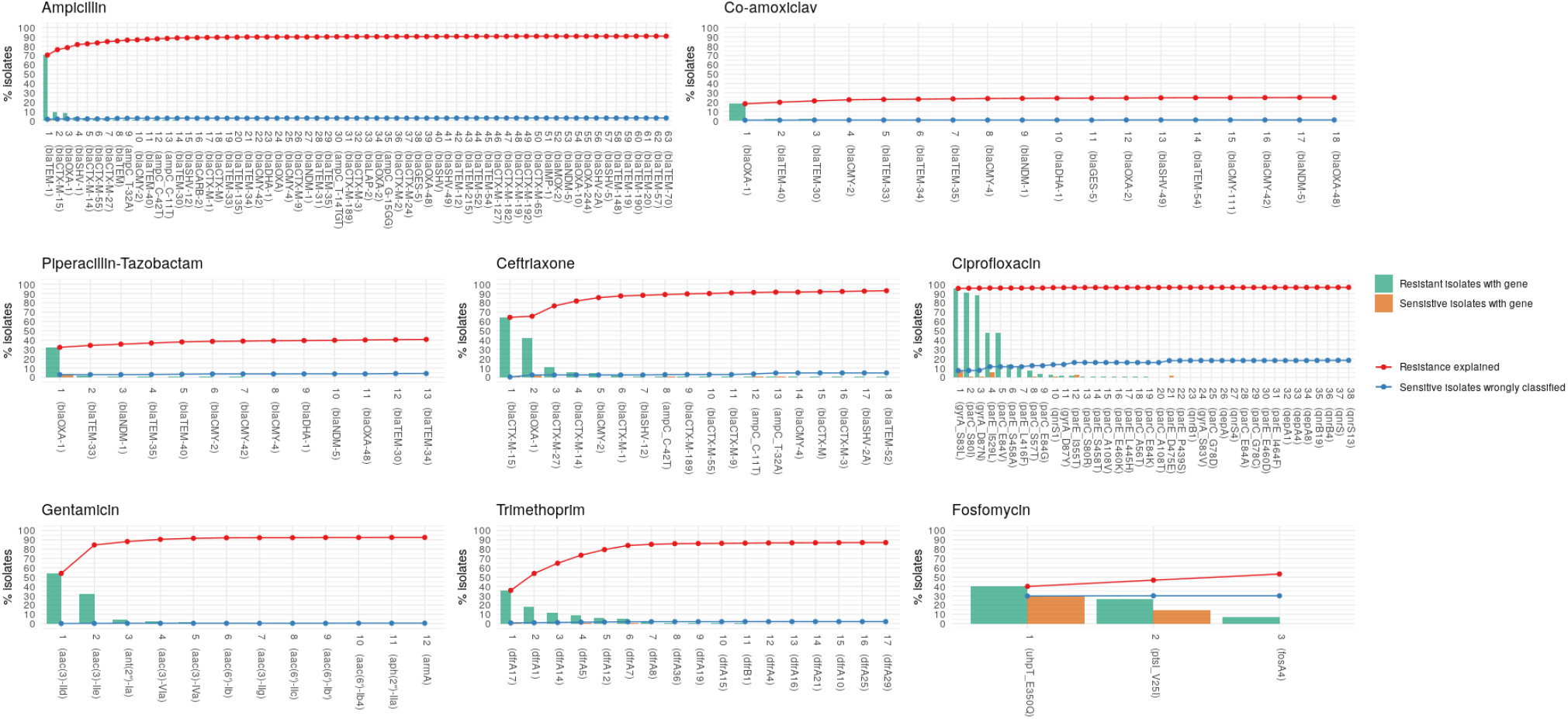
Proportion of all resistant isolates carrying known ARGs. ARGs on the x-axis are ordered by their frequency in the dataset.

### Uncatalogued variation in AMR-associated gene families

There were 7153/8586 (83.3%) isolates with at least one AMRFinder hit amongst the antibiotic classes of interest. From these we detected 161 unique AMR-associated gene families including 1292 unique ARGs within them. 158/1292 (12%) ARGs were present 10 times and 374/1292 (29%) were present 2-9 times in the dataset; of these 72/158 (46%) and 124/374 (33%) respectively had a 100% amino acid identity to the reference sequence in the AMRFinder database. The majority (760/1292 [59%]) of unique ARGs occurred only once (“singletons”; Figure 3); these could either have a limited phenotypic effect and therefore not be readily selected for, be associated with a high fitness cost and be therefore commonly lost, be currently rare (e.g. because they have recently emerged) or be bioinformatic/sequencing noise.

**Figure 3.**
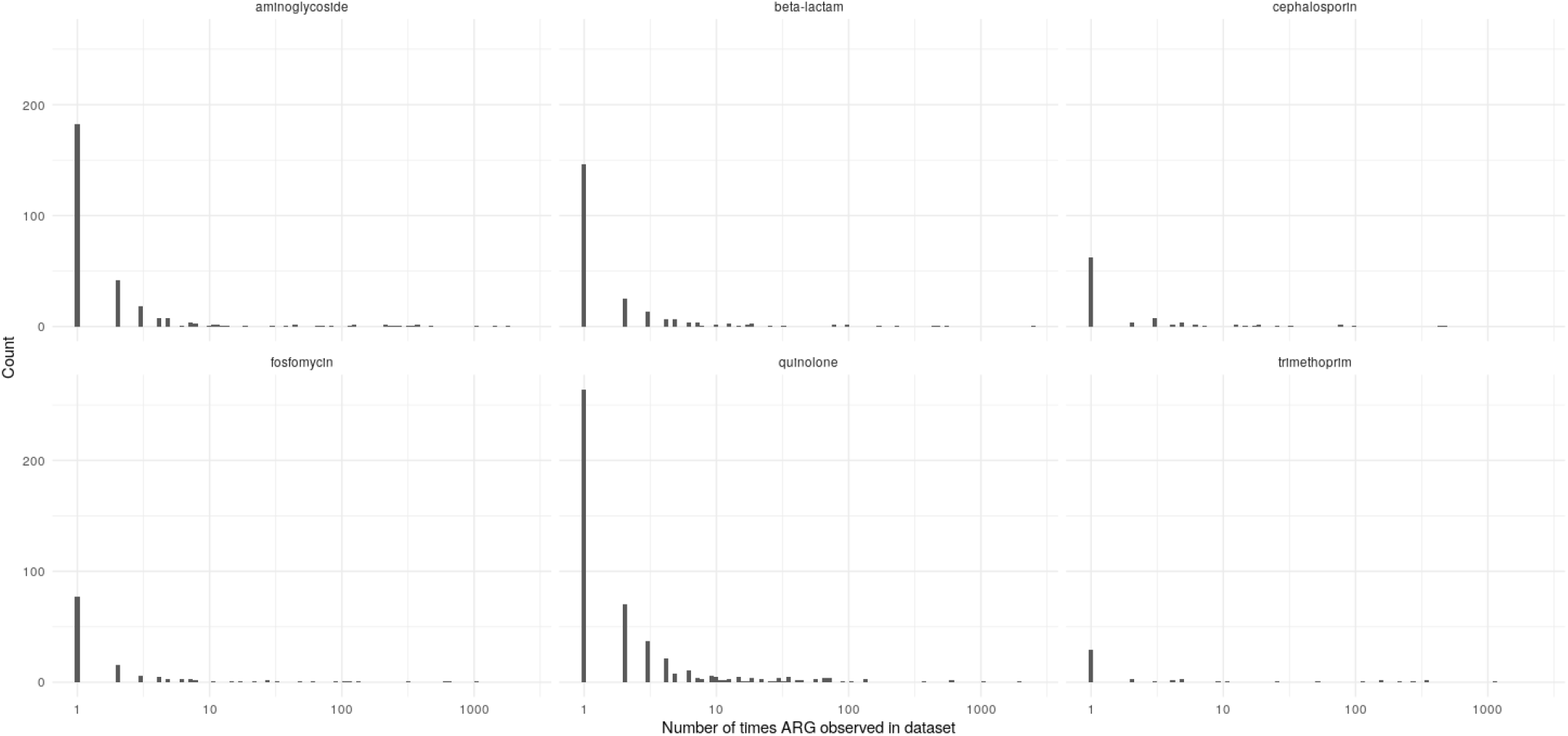
Histograms of the number of times each unique ARG is observed in the dataset, stratified by class of antibiotic.

There was no evidence of a difference in the proportion of singletons vs non-singletons which had a 100% amino acid match in the AMRFinder database (253/760 [33%] vs 197/532 [37%], p=0.18). We found similar average sequencing depths for singleton vs non-singleton genes (median 68 interquartile range IQR 51-96 vs median 69 IQR 58-101, p=0.77), suggesting that sequencing error is unlikely (Supplementary Figure 1). Whilst assembly discrepancies between SKESA and Shovill assemblies (considering a random selection of 1000 isolates with at least 1 singleton ARG) were significantly more common in singleton vs non-singleton ARGs (12/104, 11.5% vs 116/6745, 1.7%, p<0.001), the majority of singletons likely represent true background diversity rather than bioinformatic/sequencing noise.

Similar patterns of uncatalogued variation (i.e. frequent singletons, fewer examples of gene variants that appear in ≥2 isolates, and the fewest examples appearing in ≥10 isolates) in known ARGs were observed for all drugs, although for the beta-lactam, fosfomycin, quinolone and cephalosporin drugs tested, there was no evidence of a plateau in the rate of discovery of new singleton ARG alleles with increasing number of isolates (Figures 3 and 4). In contrast the accumulation curves for all drug classes plateaued when considering ARG alleles observed ≥2 times, which may be less likely to be bioinformatic/sequencing noise, in the dataset.

**Figure 4.**
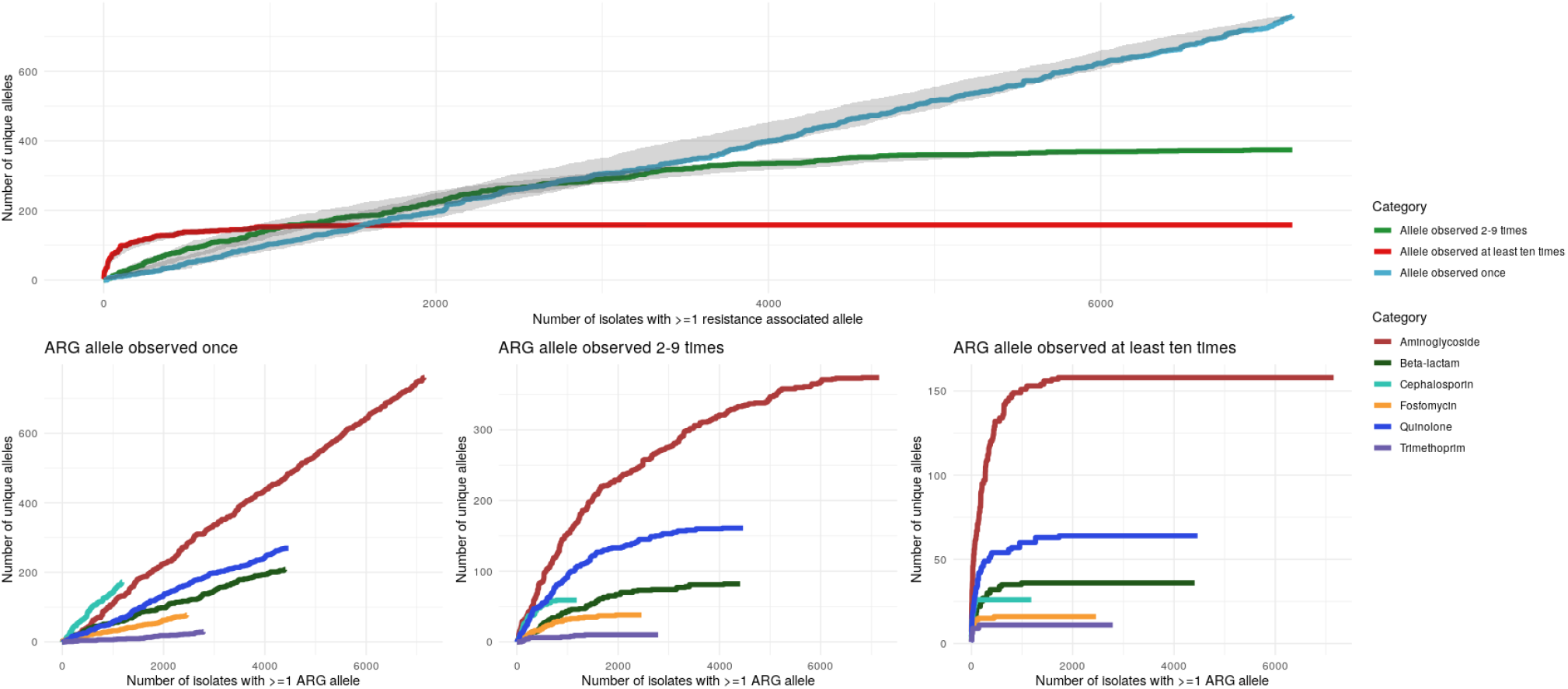
Top - simulated accumulation curves to show the relationship between the total number of unique ARGs observed across all antibiotic classes and the number of isolates with at least 1 ARG in the dataset. Coloured lines represent median estimates and with 95% confidence intervals (estimated by bootstrap approximation) shown in grey. Bottom - Simulated accumulation curves to show the relationship between the total number of unique ARGs observed and the number of isolates with at least 1 ARG of the antibiotic class denoted by the colour of the line in the dataset. Note that the Y-axis scale is different for each plot.

### Strong correlation between observed allele frequency in Oxford and globally

Overall, there was a strong relationship between the number of times an ARG allele was observed in Oxfordshire isolates and the number of times it was observed in non-Oxfordshire isolates (Spearman coefficient 0.76 (95% CI 0.72-0.79, p<0.001), Figure 5 (left)). For all drugs there were no ARG alleles observed ≥6 times in Oxfordshire isolates that were unique to this dataset (Figure 5 (right)).

**Figure 5.**
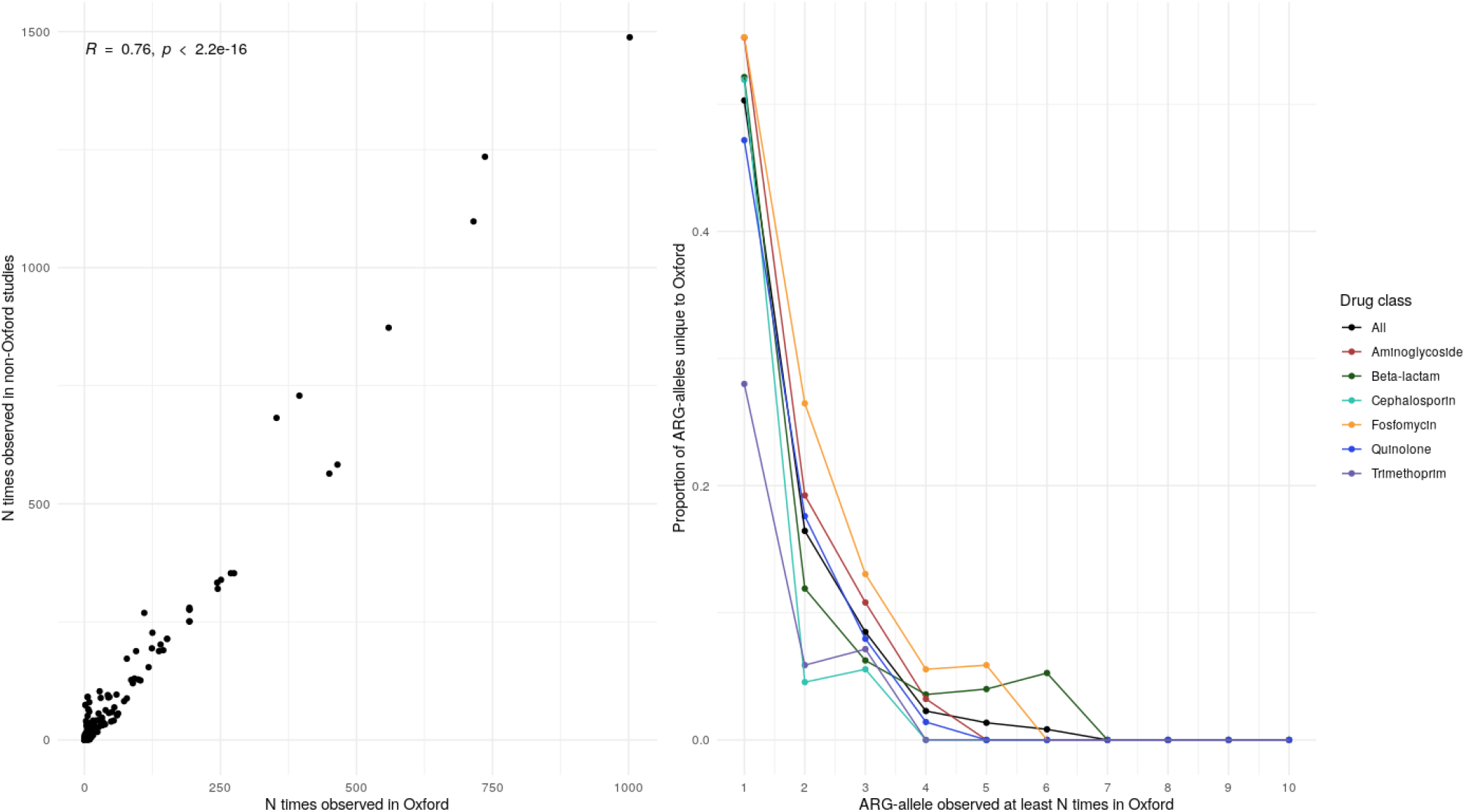
Left - Correlation between the number of times an allele was observed in Oxford and the rest of the dataset. Right - relationship between the proportion of ARG-alleles which are unique to the Oxford dataset (y-axis) compared to the number of times they are observed in the Oxford dataset (x-axis).

### Synonymous mutations associated with potentially important phenotypic differences

Using the examples of the BL-BLIs co-amoxiclav and piperacillin-tazobactam (which both currently have poorer genotype-phenotype predictive performance), we investigated whether uncatalogued variation in known AMR-associated gene families might have an important effect on AMR phenotype. There were 109 unique ARGs that clustered in the blaTEM-1 gene family, although the vast majority of sequences identified were one of four ARGs (here designated *bla*_*TEM-1 1*_ (the reference) (N=2490/3621, 69%), *bla*_*TEM-1 2*_ (95/3621, 3%), *bla*_*TEM-1 3*_ (565/3621, 16%) and *bla*_*TEM-1 4*_ (227/3621, 6%). All four had identical amino acid sequences (and hence were indistinguishable to the AMRFinder tool) but there were six synonymous polymorphic sites distinguishing these alleles (Figure 6). The remaining 244/3621 (7%) comprised 105 distinct ARGs of which 67/105 (64%)) were exact matches to known blaTEM variants (e.g. 18 *bla*_*TEM-30*_, 10 *bla*_*TEM-10*_, 7 *bla*_*TEM-34*_). After adjusting for other known alleles predicted to confer co-amoxiclav resistance (by ResFinder), there was some evidence that *bla*_*TEM-1 2*_ was associated with comparatively less resistance compared to the *bla*_*TEM-1*_ reference group (adjusted odds ratio, aOR 0.57, 95%CI 0.34-0.93, p=0.03, Table 2). Similarly, there was some evidence that *bla*_*TEM-1 3*_ was associated with reduced resistance to piperacillin-tazobactam (aOR 0.54, 95%CI 0.32-0.87, p=0.01).

**Table 2.**
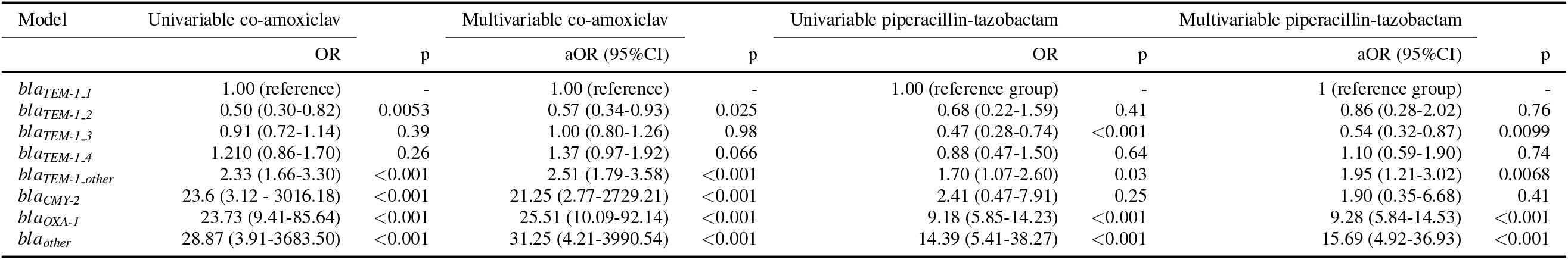
Uni- and multivariable associations with co-amoxiclav and piperacillin-tazobactam resistance. *bla*_*TEM-1 1 2 3 4*_ denote the four most common alleles of *bla*_*TEM-1*_ in the dataset (all others are denoted as *bla*_*TEM-1 other*_). *bla*_*TEM-1 1*_ is the reference version of the gene (i.e. 100% nucleotide match to the version found in the AMRFinder database). In the multivariable models, estimates are adjusted for the independent presence of the two most common BL-BLI resistance-conferring ARGs (*bla*_*CMY 2*_, *bla*_*OXA 1*_) and of any other BL-BLI ARGs grouped as *bla*_*other*_.

| Model | Univariable co-amoxiclav |  | Multivariable co-amoxiclav |  | Univariable piperacillin-tazobactam |  | Multivariable piperacillin-tazobactam |  |
| --- | --- | --- | --- | --- | --- | --- | --- | --- |
|  | OR | p | aOR (95%CI) | p | OR | p | aOR (95%CI) | p |
| *bla<sub>TEM-1.1</sub>* | 1.00 (reference) | - | 1.00 (reference) | - | 1.00 (reference group) | - | 1 (reference group) | - |
| *bla<sub>TEM-1.2</sub>* | 0.50 (0.30-0.82) | 0.0053 | 0.57 (0.34-0.93) | 0.025 | 0.68 (0.22-1.59) | 0.41 | 0.86 (0.28-2.02) | 0.76 |
| *bla<sub>TEM-1.3</sub>* | 0.91 (0.72-1.14) | 0.39 | 1.00 (0.80-1.26) | 0.98 | 0.47 (0.28-0.74) | <0.001 | 0.54 (0.32-0.87) | 0.0099 |
| *bla<sub>TEM-1.4</sub>* | 1.210 (0.86-1.70) | 0.26 | 1.37 (0.97-1.92) | 0.066 | 0.88 (0.47-1.50) | 0.64 | 1.10 (0.59-1.90) | 0.74 |
| *bla<sub>TEM-1.other</sub>* | 2.33 (1.66-3.30) | <0.001 | 2.51 (1.79-3.58) | <0.001 | 1.70 (1.07-2.60) | 0.03 | 1.95 (1.21-3.02) | 0.0068 |
| *bla<sub>CMY-2</sub>* | 23.6 (3.12 - 3016.18) | <0.001 | 21.25 (2.77-2729.21) | <0.001 | 2.41 (0.47-7.91) | 0.25 | 1.90 (0.35-6.68) | 0.41 |
| *bla<sub>OXA-1</sub>* | 23.73 (9.41-85.64) | <0.001 | 25.51 (10.09-92.14) | <0.001 | 9.18 (5.85-14.23) | <0.001 | 9.28 (5.84-14.53) | <0.001 |
| *bla<sub>other</sub>* | 28.87 (3.91-3683.50) | <0.001 | 31.25 (4.21-3990.54) | <0.001 | 14.39 (5.41-38.27) | <0.001 | 15.69 (4.92-36.93) | <0.001 |

**Figure 6.**
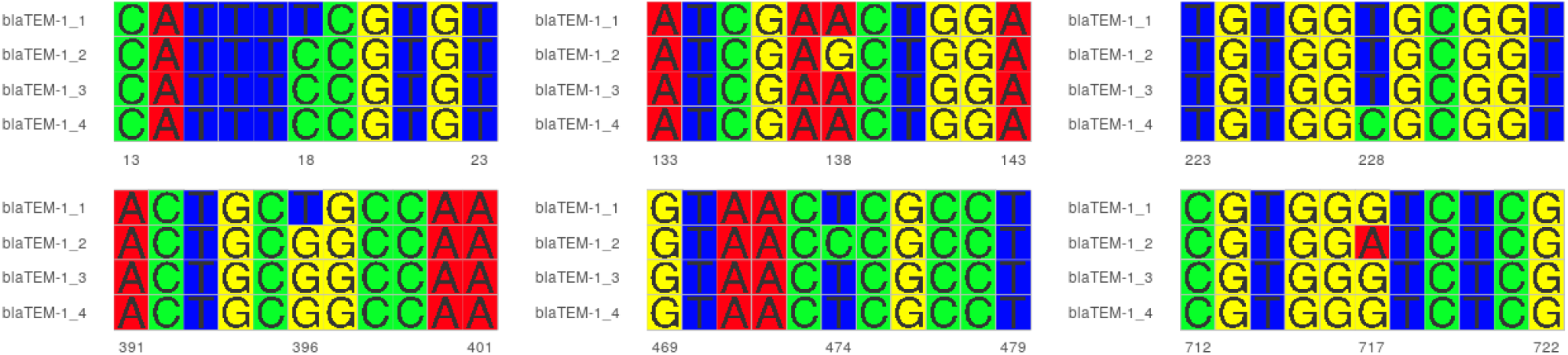
Multiple sequence alignments highlighting the six synonymous polymorphic sites identified in blaTEM-1 that distinguish the four most common alleles of this gene identified in this study.

## DISCUSSION

We analysed the ARG content of a global collection of 9001 E. coli isolates (of which 8586 had phenotypic data) to investigate how much resistance to commonly used antibiotics is explained by AMR-associated gene families included in existing catalogues and the extent of uncatalogued variation with these. For all classes of antibiotics considered here, we found that the majority of resistance is conferred by a relatively small number of ARGs. However, the fact that the performance of existing catalogues is inadequate for clinical deployment, emphasises the need for identification of more AMR-associated gene families, and/or better refinement of genotype-phenotype correlations. We showed that there is substantial background variation within known AMR-associated gene families and that better cataloguing of this (including synonymous mutations) may improve phenotypic predictions. Whilst most uncatalogued alleles were rare, those that occurred at least 6 times in the Oxford dataset were always also observed elsewhere, demonstrating strong selective pressures for convergent evolution and/or rapid global dissemination of successful genomic variation.

For all drugs considered, there was a proportion of resistance (around 10% for most drugs, higher for BL/BLIs, slightly lower for ciprofloxacin) which could not be explained by the presence/absence of known AMR-associated gene families. We hypothesise that this “sensitivity gap” is partly comprised of laboratory mistakes/mislabelling/technical failure, partly of AMR-associated gene families yet to be discovered, and partly because of phenotypic resistance which is conferred by e.g. promoter mutations, combinations of genes, and differential copy number and/or expression which are currently not effectively captured in current genotypic catalogues. The fact the most known resistance is conferred by relatively few alleles, with more numerous rarer variants explaining a much smaller proportion, suggests that very large datasets will be needed to power genome wide association studies to discover new AMR-associated gene families. An alternative approach utilising *in vitro* mutagenesis/synthetic biology may complement and speed up discovery, and enable a more refined understanding of the specific effects of mutations on resistance phenotype and fitness (i.e. growth rate)[25].

Most existing molecular epidemiology and resistance prediction studies report ARG presence/absence using default thresholds and ignore uncatalogued variation within AMR-associated gene families. This binary presence/absence approach is in contrast to the efforts made to catalogue the phenotypic effects of every mutation within AMR-associated gene families in M. tuberculosis[26], although notably the number of gene targets that are relevant to drug resistance in M. tuberculosis is much smaller than for *E. coli*. The tacit assumption of a binary presence/absence approach is that the cloud of genetic variation observed in known resistance-encoding targets is either biologically unimportant or represents artefact created by sequencing and/or bioinformatic errors and does not have an important effect on phenotype – we have shown here that this is not the case. The frequent presence of uncharacterised genetic variation with an unknown effect on phenotype is very problematic for potential clinical application of existing databases. Whilst many of the alleles detected were only observed once, there were also many that were observed frequently across datasets separated in time and space consistent with selection, and we demonstrate that such uncatalogued variation can have relevant phenotype-modifying effects and should therefore not be ignored. We hypothesise that the greater use of detailed phenotypes (i.e. using MIC as the outcome rather than a binary S/R categorisation) would further illustrate this point by providing evidence of small but potentially important incremental effects of variation on resistance. Our data also highlight that new ARG variants are being continuously generated, highlighting the potential risk that this kind of rapid genetic churn may quickly generate extended resistance phenotypes, as has been shown with blaKPC and resistance to ceftazidime-avibactam[27].

The original validation study for the AMRFinder tool measured concordance between genotypic predictions of phenotype and observed laboratory phenotype for 6,242 isolates, but only 47 of these were *E. coli*[6]. Whilst our study was not primarily designed to assess the AMRFinder tool, by evaluating its performance on a dataset of 8,586 isolates, we nevertheless have conducted by far the largest such external validation to date. None of the antibiotic classes we evaluated met the FDA criteria for acceptable maj and vmj discrepancy rates. In addition to the possible improvements described above, more granular drug-level classification of ARGs should be a priority for the AMRFinder tool and would likely improve predictive performance (highlighted by the overall slightly better performance of ResFinder in a recent validation study[10] and by better performance of existing tools when using curated gene-drug associations[9, 28]). Our study also highlights potential phenotypic differences for alleles of AMR-associated gene families which differ by synonymous mutations, suggesting that classification of ARGs using amino-acid sequences alone should be avoided.

Limitations of this study include the fact that the dataset over-represents European clinical isolates which may reflect a different picture from AMR gene diversity and selection in other potential niches/geographical regions. Whilst all included studies used EUCAST breakpoints, it is possible that technical laboratory-level differences in standard operating procedures may explain some of the variation of phenotypes observed. Some of the variation in resistance to BL-BLI (and possibly other) antibiotics may be explained by considering other factors related to ARG presence e.g. copy number variation in blaTEM-1/promoter mutations which we did not explore here[11].

In summary, we demonstrate substantial variation in known AMR gene targets in *E. coli*, and that some of these variants are selected across space and time. Surveillance approaches taking this hitherto uncatalogued variation into account might be able to more rapidly identify genetic variants that are emerging/disseminating. We also highlight three areas of focus for the improvement of existing ARG databases. For most drug classes, current knowledge explains most, but not all, resistance and so new gene target discovery either by combining large global sequencing/and detailed phenotyping efforts to power discovery of rare/small effect size AMR-associated gene families or *in vitro* mutagenesis studies characterising these genes and the effects of genetic variation are needed. For new, as yet undiscovered AMR-associated gene families, it will be important to develop rules for systematically cataloguing new alleles so that their phenotypic effect can be properly considered. Finally, the application of databases needs to be improved to consider mutations at both the nucleotide and amino-acid level as well as the effects of these changes on phenotypes at the specific drug-species level.

## Data Availability

All assemblies are available at 10.6084/m9.figshare.22220212 and associated metadata can be found in supplementary dataset 1. All code used for the analysis can be found at https://github.com/samlipworth/resistome_variation where there is also a binder environment in which the key aspects of the analysis can be replicated.

## ACKNOWLEDGMENTS

We would like to thank the authors of the datasets used in this study for making their data freely available for public use. The computational aspects of this research were funded from the NIHR Oxford BRC with additional support from the Wellcome Trust Core Award Grant Number 203141/Z/16/Z. SL was funded by an MRC Clinical Research Training Fellowship MR/T001151/1. ASW and TEAP are also supported by the NIHR Oxford Biomedical Research Centre. ASW is an NIHR Senior Investigator. NS is an NIHR Oxford BRC Senior Fellow. This research is supported by the National Institute for Health Research (NIHR) Health Protection Research Unit in Healthcare Associated Infections and Antimicrobial Resistance (NIHR200915), a partnership between the UK Health Security Agency (UKHSA) and the University of Oxford. The views expressed are those of the author(s) and not necessarily those of the NIHR, UKHSA or the Department of Health and Social Care. This research was supported by the National Institute for Health Research (NIHR) Oxford Biomedical Research Centre (BRC). The views expressed are those of the author(s) and not necessarily those of the NHS, the NIHR or the Department of Health.

