## Supplementary figures and images for "Exploring the extent of uncatalogued genetic variation in antimicrobial resistance gene families in *Escherichia coli*"

### Supplementary figure

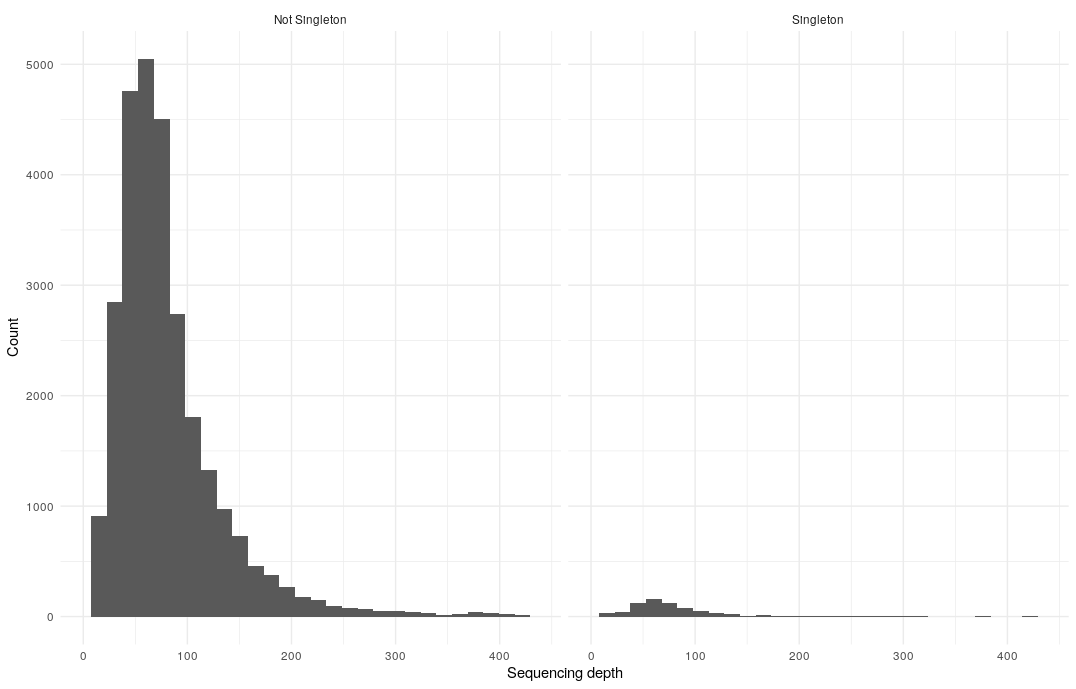

Figure S1: Distribution of sequencing depths for singleton vs non-singleton genes
